# Analysis of factors influencing patient delay by patients with pulmonary tuberculosis in HuYanghe City, Xinjiang Province

**DOI:** 10.64898/2026.01.16.26344265

**Authors:** Fukui He, Xin Wang, Kaihao Wang, Junxia Yan

## Abstract

**Objective:** The purpose of this study was to systematically describe the current status of patient delay among pulmonary tuberculosis (TB) patients in Huyanghe City, Xinjiang, from 2014 to 2024, and to examine factors associated with patient delay, thereby providing an evidence base for developing targeted interventions to reduce delays in care seeking and strengthen TB prevention and control.

**Methods:** This observational study was based on the Tuberculosis Patient Management Subsystem of the China CDC information system. We extracted patient delay information for patients with pulmonary tuberculosis registered and reported by TB-designated hospitals in Huyanghe City from 2014 to 2024. Continuous variables were summarized using the median and interquartile range (IQR). Factors associated with patient delay were explored using univariable analyses and multivariable logistic regression models.

**Results:** Between 2014 and 2024, TB-designated facilities in Huyanghe City registered 1,021 pulmonary TB patients, and 534 had patient delay (52.00%).Patient delay lasted a median of 24 days (IQR, 9~62).In multivariable models, male sex (OR:0.702, 95% CI:0.537~0.917), age>60 years (OR:0.759, 95%CI:0.584~0.985), and presentation during the COVID-19 period (OR:0.658, 95%CI:0.465~0.932) were linked to reduced odds of delay. Conversely, Han ethnicity. (OR:1.936, 95% CI:1.168~3.207), pathogen positive (OR:1.923, 95% CI:1.079~3.426), comorbidities conditions (OR:1.652, 95% CI:0.987~2.764), and Employed (OR:2.293, 95% CI:1.694~3.103) increased the odds of delay.

**Conclusions:** Patient delay in pulmonary TB remained high in Huyanghe City from 2014 to 2024, yet declined overall. Younger age (<60 years), female, comorbidity, employment, and microbiological positivity may predispose pulmonary TB patients in this setting to delayed care seeking.

---

Tuberculosis(TB), caused by Mycobacterium tuberculosis and transmitted mainly through the airways, most often manifests as pulmonary TB and remains a major infectious threat in China.In China, the estimated number of incident TB cases in 2024 was 696,000 (741,000 in 2023), with an estimated incidence of 49 per 100,000 population, representing a 5.8% decrease compared with 2023; among the 30 high-burden TB countries, China’s estimated TB incidence ranking decreased from third to fourth, accounting for 6.5% of global incident cases ^**[1]**^.Prior evidence suggests substantial patient delay in pulmonary TB ^**[2]**^, with some patients not seeking care at all ^**[3]**^. A global meta-analysis showed wide variation in patient delay, with median values spanning 4 to 199 days; ~42% of pulmonary TB patients first sought care ≥ 30 days after symptom onset, and reported medians ranged 2 to 128.5 days (IQR:12~34), underscoring marked regional heterogeneity ^**[4]**^. In China, surveillance data indicate longer patient and overall delays in local residents than in migrants; among older adults, women are more prone to patient delay, whereas lower education is associated with shorter overall delay ^**[5]**^. In a Dalian, China cohort of 18,100 incident pulmonary TB cases, the median time to treatment initiation was 30 days (IQR, 14~59); delays >53 days predicted unfavourable outcomes and >103 days predicted mortality, supporting their use as operational risk thresholds ^**[6]**^.Meta-analytic evidence links longer patient delay to low education/health literacy, rural residence, female sex, poverty, stigma and misconceptions, and alcohol use.Longer delay has also been linked to older age, migrant status, subtle or atypical symptoms, chronic cough, and system-level barriers, including non-specialist first contact, poor referral, repeated antibiotic-treated visits without TB work-up, smear negativity, extrapulmonary TB, and limited diagnostic capacity.In contrast, haemoptysis, chest pain, and multiple symptoms tend to accelerate care seeking; rapid molecular testing (e.g., Xpert) and streamlined referral and laboratory pathways may shorten delays ^**[7]**^, and randomized trials suggest that integrating traditional care with TB programmes, alongside provider training and referral collaboration, can reduce both patient and diagnostic delays ^**[8]**^.Health education, higher education levels, and active community screening are linked to shorter patient delay ^**[4,8]**^, but estimates vary widely across studies ^**[4]**^.

Xinjiang ranks among the highest in China for notified pulmonary TB, underscoring major prevention and control challenges ^**[9]**^.Recent evidence indicates that older pulmonary TB patients in Xinjiang had an overall diagnostic delay rate of 18.66% during 2018 – 2024, with higher patient delay among farmers and residents of northern Xinjiang division-level cities ^**[10]**^.Located at the centre of northern Xinjiang’s triangular economic zone, Huyanghe combines high connectivity and population mobility with rapid urbanization and a sizeable agricultural population, creating a complex landscape of determinants for TB care seeking and diagnosis.Yet empirical evidence on the scale, risk patterns, and drivers of TB patient delay in Huyanghe and neighbouring northern Xinjiang division-level cities remains scarce, limiting locally tailored control efforts.Systematic investigation is needed to quantify delays and pinpoint high-risk groups and modifiable determinants.Using 2014-2024 data from Huyanghe, we characterize patient delay and its determinants, informing analyses of patient delay in northern Xinjiang and supporting patient-pathway-guided interventions for earlier detection, diagnosis, and treatment.

## Study patients and methods

### Study patients and setting

We extracted records of 1,030 pulmonary TB patients with current residence in Huyanghe City who were registered from January 1, 2014 to December 31, 2024 in the China CDC system, capturing demographics, residence, registration year, symptom onset and first-visit dates, case source, treatment category, diagnostic classification, key-population status, and comorbidities.We excluded cases classified as NTM co-infection, extrapulmonary TB, or with implausible symptom-onset and first-visit dates, yielding 1,021 pulmonary TB records for analysis.

Relevant definitions

Pulmonary TB diagnosis and classification:We followed Chinese standards (WS 288 – 2017; WS 196 – 2017), and pulmonary TB in this study encompassed tuberculous pleurisy.

Definitions:We used commonly applied Chinese definitions ^[11]^: patient delay (days), the interval from the onset of any TB-suggestive symptom to the first healthcare visit ^[2]^; patient delay, defined as >14 days ^[12]^;patient delay rate, calculated as delayed cases divided by all registered pulmonary TB cases ×100% ^[13]^.

Initial treatment:Did not previously receive anti-TB treatment or had been treated for less than one month.

Retreatment: Previously received anti-TB treatment for more than one month.

Etiology result:Results of the M. tuberculosis test using sputum smear microscopy, PCR, or culture. A”positive”result indicates the detection of M. tuberculosis, while a”negative” result indicates that M. tuberculosis was not detected.

Comorbidities: HIV co-infection or diabetes diagnosed before or concurrent with TB.

Patient source:The type of setting that the patient came from.

Transfer:Patients were transferred from non-TB-designated medical institutions or non-TB outpatient clinics.

Referral:Patients were referred by doctors at primary healthcare institutions due to symptoms similar to pulmonary TB or with radiological evidence suggesting suspected pulmonary TB.

### Statistical analysis

Data were curated in Excel 2016 and analysed in R (v4.3.2).Non-normally distributed continuous variables are reported as median (M; Q25–Q75) and categorical variables as n (%). Chi-squared tests or Fisher’s exact probability method were utilized for univariate analysis of the factors influencing the patient delay by patients with pulmonary TB, while the logistic regression model was utilized for multivariate analysis. *P*<0.05 is statistically significant.

## Results

### Patterns of patient delay in pulmonary tuberculosis

From 2014 to 2024, 1,030 TB cases were registered at TB-designated facilities in Huyanghe City; after excluding implausible or incomplete records, 1,021 were retained (99.13% included).The sample included 623 men (61.02%) and 398 women (38.98%), for a male-to-female ratio of 1.57:1.The 45 to <65-year group predominated (603/1,021; 59.59%).Han ethnicity accounted for 946 patients (92.65%).Most were non-employed (homemakers/unemployed or other; 756/1,021, 74.05%).Most patients were detected via passive case finding (857/1,021; 83.94%).Comorbidity information was unavailable for 761 patients (74.53%) (**Table 1**).

**Table 1.**
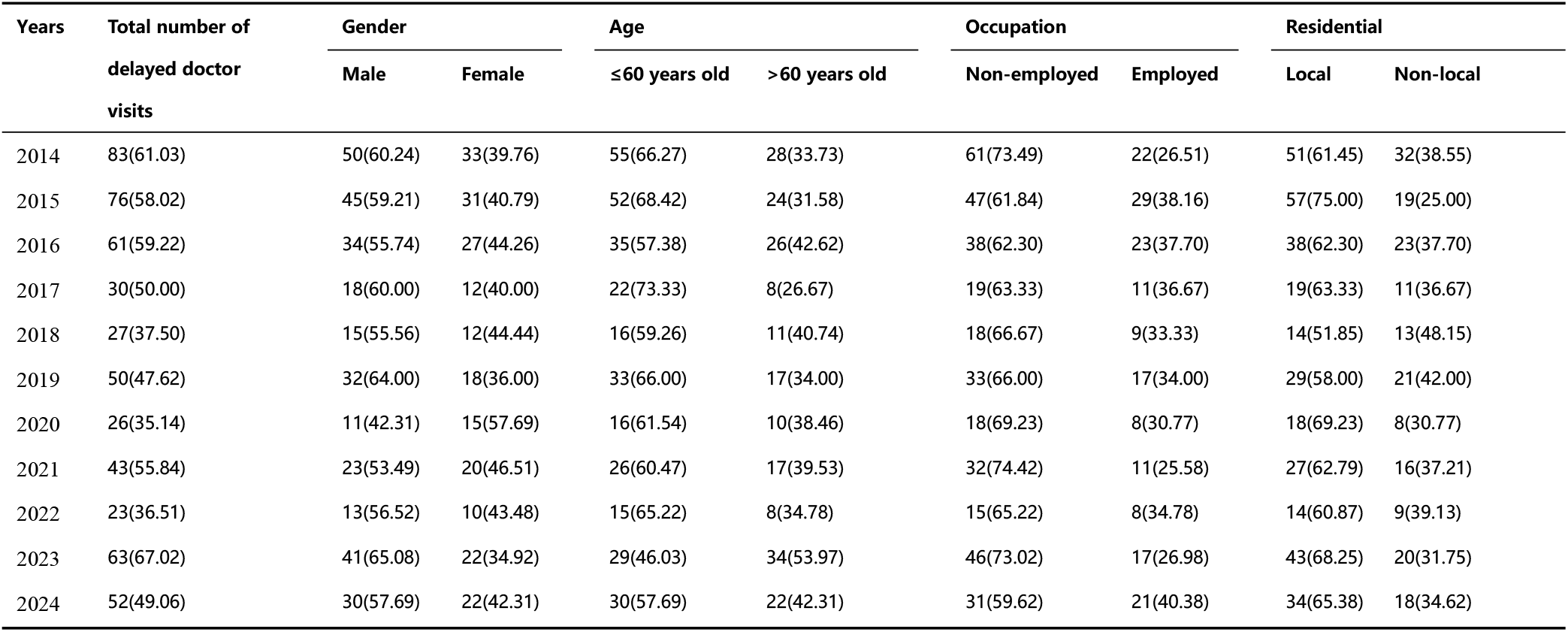
Patient delay by patients with TB from 2014 to 2024(%)

### Patient delay among patients with pulmonary tuberculosis

Over 2014–2024, the patient delay rate declined with modest fluctuations (χ^2^ _trend_= 3.87, *P* = 0.05), from 61.02% in 2014 to 49.06% in 2024., the patient delay rate declined with modest fluctuations (χ^2^_trend_ = 3.87, *P* = 0.05), from 61.02% in 2014 to 49.06% in 2024.A similar downward pattern was observed in men (χ^2^_trend_= 3.193, *P* = 0.074) and in the 45 to <65-year group (χ^2^_trend_ = 5.705, *P* = 0.017).

In 2014–2024, the median time from symptom onset to first visit among 1,021 pulmonary TB patients in Huyanghe City was 15 days (IQR, 6–40); 534 (52.00%) met criteria for patient delay, with a median delay of 24 days (IQR, 9–62). Delays clustered at 1–29 days (295, 55.24%), 30–59 (102, 19.10%), 60–89 (59, 11.05%), 90–179 (61, 11.42%), and ≥180 days (17, 3.18%). In univariable analyses, delay rates varied significantly by ethnicity, township, bacteriological status, comorbidity, the COVID-19 period, and occupation (all *P* < 0.05) **(Table 2)**.

**Table 2.** Patient delay among tuberculosis patients, 2014–2024.

| Variable | Total patients. | Time to first visit | Delay duration | Non-delayed patients | Delayed patients | $\chi^2$ | $P$ |
| --- | --- | --- | --- | --- | --- | --- | --- |
|  | [Case(Proportion, %)] | [d, M(Q25, Q75)]<sup>a</sup> | [d, M(Q25, Q75)]<sup>a</sup> | [Case(Proportion, %)] | [Case(Proportion, %)] |  |  |
| Age (years old) |  |  |  |  |  | 3.23 | 0.091 |
| ≤60 | 603 (59.06) | 17 (6, 47) | 27 (11, 62) | 329 (54.56) | 274 (45.44) |  |  |
| >60 | 418 (40.94) | 14 (5, 31) | 18 (7, 58) | 205 (48.92) | 213 (51.08) |  |  |
| Gender |  |  |  |  |  | 3.16 | 0.080 |
| Female | 398 (38.98) | 17 (7, 41) | 22 (8, 44) | 176 (44.22) | 222 (55.78) |  |  |
| Male | 623 (61.02) | 15 (4, 40) | 26 (9, 69) | 311 (49.92) | 312 (50.08) |  |  |
| **Ethnicity** |  |  |  |  |  | 7.27 | <0.012 |
| Non-Han | 75 (7.35) | 9 (1, 26) | 18 (8, 35) | 47 (62.67) | 28 (37.33) |  |  |
| Han . | 946 (92.65) | 16 (6, 43) | 25 (9, 62) | 440 (46.51) | 506 (53.49) |  |  |
| **Key population.** |  |  |  |  |  | 1.41 | 0.241 |
| No | 831 (81.39) | 16 (6, 37) | 21 (8, 55) | 389 (46.81) | 442 (53.19) |  |  |
| Yes | 190 (18.61) | 14 (2, 53) | 41 (19, 81) | 98 (51.58) | 92 (48.42) |  |  |
| **Residential** |  |  |  |  |  | 0.84 | 0.360 |
| Local | 671 (65.72) | 15 (5, 39) | 24 (8, 59) | 327 (48.73) | 344 (51.27) |  |  |
| Non-local | 350 (34.28) | 17 (7, 44) | 24 (11, 64) | 160 (45.71) | 190 (54.29) |  |  |
| **TB-suggestive symptoms** |  |  |  |  |  | 1.01 | 0.321 |
| None /unspecified | 761 (74.53) | 16 (6, 41) | 24 (8, 62) | 356 (46.78) | 405 (53.22) |  |  |
| Yes | 260 (25.47) | 14 (5, 37) | 24 (9, 58) | 131 (50.38) | 129 (49.62) |  |  |
| **Patient source** |  |  |  |  |  | 0.22 | 0.640 |
| Passive | 857 (83.94) | 15 (6, 42) | 25 (10, 64) | 406 (47.37) | 451 (52.63) |  |  |
| Proactive | 164 (16.06) | 15 (2, 34) | 20 (6, 44) | 81 (49.39) | 83 (50.61) |  |  |
| **Pathogenic results** |  |  |  |  |  | 21.93 | <0.011 |
| Positive | 405 (39.67) | 18 (5, 43) | 24 (8, 58) | 173 (42.72) | 232 (57.28) |  |  |
| Negative | 441 (43.19) | 17 (6, 46) | 27 (10, 69) | 203 (46.03) | 238 (53.97) |  |  |
| Tuberculous pleurisy | 114 (11.17) | 11 (4, 23) | 22 (8, 43) | 73 (64.04) | 41 (35.96) |  |  |
| No bacteriological results | 61 (5.97) | 10 (2, 25) | 15 (9, 38) | 38 (62.30) | 23 (37.70) |  |  |
| **Comorbidities** |  |  |  | (0.00) |  | 4.44 | 0.040 |
| No | 946 (92.65) | 15 (6, 41) | 26 (10, 64) | 460 (48.63) | 486 (51.37) |  |  |
| Yes | 75 (7.35) | 19 (6, 31) | 12 (6, 38) | 27 (36.00) | 48 (64.00) |  |  |
| **Initial/retreatmen** |  |  |  |  |  | 2.34 | 0.131 |
| Retreatment | 90 (8.81) | 22 (7, 42) | 22 (15, 48) | 36 (40.00) | 54 (60.00) |  |  |
| Initial treatmen | 931 (91.19) | 15 (6, 40) | 24 (8, 63) | 451 (48.44) | 480 (51.56) |  |  |
| **During the COVID-19 period.** |  |  |  |  |  | 9.87 | <0.010 |
| Pre – COVID-19 period | 601 (58.86) | 17 (6, 46) | 28 (10, 62) | 276 (45.92) | 325 (54.08) |  |  |
| COVID-19 period. | 222 (21.74) | 12 (4, 31) | 21 (10, 57) | 126 (56.76) | 96 (43.24) |  |  |
| Post – COVID-19 period | 198 (19.39) | 17 (6, 39) | 17 (6, 52) | 85 (42.93) | 113 (57.07) |  |  |
| **Occupation** |  |  |  |  |  | 28.58 | <0.010 |
| Non-employed | 756 (74.05) | 14 (6, 27) | 15 (5, 28) | 398 (52.65) | 358 (47.35) |  |  |
| Employed | 265 (25.95) | 65 (1, 107) | 78 (52, 124) | 89 (33.58) | 176 (66.42) |  |  |

**Note** \*: Values are reported as whole numbers after rounding. Patient source: Cases were categorized as active case finding (health examinations, targeted screening, school-entry screening, close-contact screening) or passive detection (symptom-driven presentation, transfer/referral, provider referral, tracing, or other); TB-suggestive symptoms: Respiratory symptoms (cough, sputum, dyspnoea, chest pain, etc.); “no suggestive symptoms” was defined as documented asymptomatic status or no symptom entry in the record. Comorbidities: HIV positivity or diabetes versus none/unknown. COVID-19 period: pre-COVID (2017 – 2019), COVID period (2020 – 2022), and post-COVID (2023), based on year of registration. Key population: Membership in priority groups (school/childcare staff, supervised populations, or close contacts of active pulmonary TB cases).

### Multivariate logistic regression analysis

We fitted a multivariable logistic regression model including sex, age group, ethnicity, comorbidity, treatment category, COVID-19 period, and occupation, with patient delay as the outcome.The results showed that, compared with patients aged <60 years, male patients (OR: 0.702, 95% CI: 0.537–0.917), those aged ≥60 years (OR: 0.759, 95% CI: 0.584– 0.985), and those presenting during the COVID-19 pandemic period (OR: 0.658, 95% CI: 0.465 – 0.932) had significantly lower odds of patient delay. Conversely, non-minority ethnicity (OR: 1.936, 95% CI: 1.168 – 3.207), bacteriological positivity (OR: 1.923, 95% CI: 1.079 – 3.426), comorbidity (OR: 1.652, 95% CI: 0.987 – 2.764), and employment (OR: 2.293, 95% CI: 1.694–3.103) increased the odds of delay(**Table 3**).

**Table 3.** Multivariate logistic regression analysis of factors influencing patient delay by patients with TB.

| Variable | $\beta$ | *S.E* | *Z* | *P* | *OR* | 95%*CI* |
| --- | --- | --- | --- | --- | --- | --- |
| Intercept | -0.826 | 0.399 | -2.070 | 0.038 | 0.438 | 0.200 ~ 0.957 |
| Male | -0.354 | 0.136 | -2.600 | 0.009 | 0.702 | 0.537 ~ 0.917 |
| Age>60 years old | -0.276 | 0.133 | -2.071 | 0.038 | 0.759 | 0.584 ~ 0.985 |
| Han Ethnicity | 0.660 | 0.258 | 2.563 | 0.010 | 1.936 | 1.168 ~ 3.207 |
| Pathogenic results |  |  |  |  |  |  |
| Tuberculous pleurisy | -0.198 | 0.342 | -0.579 | 0.563 | 0.820 | 0.419 ~ 1.604 |
| Negative | 0.495 | 0.305 | 1.626 | 0.104 | 1.641 | 0.903 ~ 2.981 |
| Positive | 0.654 | 0.295 | 2.219 | 0.026 | 1.923 | 1.079 ~ 3.426 |
| Comorbidities (Yes) | 0.502 | 0.263 | 1.910 | 0.056 | 1.652 | 0.987 ~ 2.764 |
| During the COVID-19 period |  |  |  |  |  |  |
| COVID-19 period | -0.418 | 0.177 | -2.358 | 0.018 | 0.658 | 0.465 ~ 0.932 |
| Post-COVID-19 period | 0.155 | 0.192 | 0.806 | 0.420 | 1.168 | 0.801 ~ 1.702 |
| Employed | 0.830 | 0.154 | 5.374 | <.001 | 2.293 | 1.694 ~ 3.103 |

## Discussion

We observed a median time to first visit of 15 days (IQR, 6–40) in Huyanghe City, with a delay rate of 52.00%, similar to Huzhou (52.9%) ^**[14]**^ and above the national level (47.38%) ^**[15]**^, yet below Yantai (68.5%) ^**[16]**^, Urumqi (68.06%) ^**[17]**^, Qinghai (59.60%) [18], and Dongguan (60.3%) ^**[19]**^.It exceeded reported rates in Shanghai (37.89%) [20], Yinchuan (46.54%) ^**[21]**^, and Beijing’s Chaoyang District (42.60%) ^**[22]**^.The median patient delay was 24 days (IQR, 9–62), similar to the 24.6 days reported in Ethiopia ^**[16]**^, higher than that in Qinghai Province (21 days) [18], and lower than those reported in Hanzhong (26 days) ^**[23]**^, Heilongjiang Province (35 days) [24], Mongolia (28 days) ^**[25]**^, and Lishui (36 days) ^**[26]**^; internationally, it was also lower than estimates from Montenegro (30 days) ^**[27]**^ and Switzerland (36 days) ^**[28]**^.Notably, delays showed a clear long-tail distribution, suggesting that many patients remained in the community for extended periods and hesitated to seek care after symptoms began.

Over 2014–2024, patient delay in Huyanghe City declined overall, from 61.02% in 2014 to 49.06% in 2024, despite modest year-to-year fluctuations.This decline may reflect cumulative gains in TB services: standardized tiered and designated care with clearer referral pathways, coupled with routine health education, may curb post-symptom “wait-and-see” behaviour and self-medication, shortening delays.Expanded diagnostic capacity, including rapid molecular testing, may reduce opportunity costs and promote earlier presentation ^**[29]**^.Strengthened active screening and key-population management may move case finding upstream ^**[30]**^, reducing untreated community time and compressing the transmission window. The COVID-19 pandemic (from 2019) may also have contributed [31,32], as heightened sensitivity to respiratory symptoms and the expansion of fever-clinic screening likely lowered the threshold for evaluation of cough or fever, shortening time to first visit. In line with this, diagnostic delay during COVID-19 was driven mainly by transport restrictions rather than personal factors, and only 3.4% of 3,224 patients on anti-TB therapy reported irregular adherence ^**[33]**^.Nevertheless, follow-up examinations and treatment during the course of TB care may still have been affected by the pandemic ^**[34,35]**^.

Sociodemographic correlates of lower delay included age ⩾ 60 years, male sex, and non-minority ethnicity. The lower delay among older adults aligns with prior reports ^**[36,37]**^. In contrast to Xu et al. ^**[38]**^ and Guo et al. ^**[26]**^, older patients (>60 years) in Huyanghe had a lower delay rate (51.08%) than those reported in Dongguan (74.3%) ^**[39]**^ and Lishui (76.6%) ^**[26]**^, potentially reflecting expanded routine examinations and targeted TB screening in older and other priority groups that facilitate earlier referral once symptoms emerge.Context may also matter: with many older residents and high mobility in a largely agricultural population, time costs are salient, and early symptoms may prompt self-medication or first contact with primary care or pharmacies rather than TB-designated hospitals. Second, cross-regional mobility may limit awareness of local designated TB services, leading to more dispersed choices of first-contact providers and longer referral chains, thereby prolonging the time to the first standardized healthcare visit ^**[40]**^. In contrast, older long-term residents may be detected earlier through routine family-doctor follow-up and periodic health checks for chronic disease management.In our study, among 350 non-local patients, the patient delay rate was 54.29%; although the difference was not statistically significant in univariable analyses, the quarterly follow-up typically provided by designated clinicians at local community health centers for chronic conditions such as diabetes and hypertension may, to some extent, increase opportunities for early identification.

Male sex was associated with a lower risk of patient delay. This pattern accords with prior evidence on sex differences. Storla and colleagues noted that female sex is an important contributor to diagnostic and treatment delays in pulmonary tuberculosis, and its impact is often amplified by psychosocial factors and limited access to health services; caregiving responsibilities, financial and time constraints, and disease-related stigma may all postpone women’s initial healthcare seeking ^**[41,42]**^.Evidence from Chinese populations also supports this conclusion: women often have lower socioeconomic status and are more likely to delay care seeking after TB-related symptoms occur ^**[43,44]**^.In addition,Sex-related biological differences may yield milder symptomatology in women than in men, potentially promoting delay ^**[45]**^. By contrast, non-minority patients constitute a large proportion of the local population and show greater heterogeneity in occupation and mobility; consequently, their care-seeking pathways may be more dispersed, with more frequent first contact at non–TB-designated facilities or primary care sites and a higher likelihood of self-medication. Moreover, the sample size of ethnic minority participants in this study was relatively small, and we were unable to perform stratified analyses by potential confounders such as educational attainment, health literacy, and economic status; future studies in multi-ethnic settings should incorporate residential patterns, primary-care coverage, and health-behaviour profiles to enable stratified analyses and more accurately identify populations at high risk of delayed presentation. We observed higher delay among bacteriologically positive cases than among those without microbiological results; given their prolonged infectiousness and symptom progression from mild to severe, diagnostic delay in this group is especially consequential ^**[46]**^.In this study, the delay rate among bacteriologically positive pulmonary TB patients (57.28%) was higher than that among patients with no bacteriological results (37.70%) and those who were bacteriologically negative (53.97%), indicating bacteriological positivity as a risk factor for patient delay. These rates remained lower than those reported for bacteriologically positive older pulmonary TB patients in Lishui (73.22%) [26] and Dongguan (81.11%) ^**[39]**^.

Delay tended to be higher among patients with comorbidity and among those in employment; 75 of 1,021 patients had comorbidities, and their delay rate was 64.00%, in line with prior findings ^**[47]**^. Some studies have reported a lower delay rate among pulmonary TB patients with diabetes than among those without diabetes ^**[26]**^, possibly because TB patients with diabetes may present with more overt clinical manifestations, more extensive pulmonary lesions on imaging, and higher bacteriological positivity, thereby prompting earlier care seeking. However, the pattern may be reversed among HIV-positive patients, as immunosuppression may delay the onset of TB-suggestive symptoms, making them less perceptible and reducing timely care seeking. Multiple studies have also shown that pulmonary TB patients with chronic comorbidities (e.g., cardiovascular disease) are more likely to attribute non-specific symptoms such as cough, fatigue, and weight loss to their pre-existing conditions, thereby lowering vigilance for TB and delaying the first healthcare visit ^**[5,48]**^. Moreover, being employed was associated with a higher risk of patient delay and showed the largest effect size in the multivariable analysis. This association may be related to time costs, financial pressures, and work-related constraints ^**[42]**^. Studies in China and other low- and middle-income countries have repeatedly identified agricultural workers and migrant laborers as high-risk groups for delayed care seeking, characterized by limited access to healthcare, poor awareness of TB-designated services, and inefficient referral after first contact with primary or private facilities ^**[5,49]**^. An earlier systematic review by Storla et al. highlighted that first presentation to non-TB specialist services followed by repeated symptomatic treatment is a core mechanism underlying patient delay, and this pathway is more common among employed populations ^**[50]**^. These findings suggest that reducing patient-side delays will require supply-side improvements and cross-regional medical coverage, thereby mitigating structural barriers to care seeking among employed populations arising from work constraints.

A key limitation is that we restricted inclusion to cases captured in the TB Management subsystem of the China CDC information system. Although the sample size was relatively large, the findings may not be generalizable to the entire population.We also lacked data on several potentially important determinants, including socioeconomic status, education, insurance coverage, behavioural and psychological factors, and TB knowledge. During data processing, discrepancies between healthcare providers and patients in interpreting the time of first onset of TB-suggestive symptoms, together with substantial missing symptom documentation in medical records, may have led to misclassification of patient delay. Targeted surveys are needed to better capture and explain determinants of care-seeking behaviour. Furthermore, intervention studies addressing diagnostic delay in tuberculosis and evaluations of their effectiveness remain limited; patient-pathway–oriented intervention research is urgently needed to test the real-world effectiveness of different strategies in reducing both patient and diagnostic delays.

## Conclusion

In summary, the patient delay rate for pulmonary tuberculosis in Huyanghe City remained high during 2014–2024, indicating a persistently concerning situation; although the overall delay rate has declined in recent years, a slight rebound has been observed since 2021, underscoring the urgent need to strengthen active screening and comprehensive control measures for priority groups, including individuals aged <60 years, those with comorbidities, and mobile populations. Role-tailored health education should be delivered to different target populations, promoting routine health examinations and proactive screening, encouraging individuals with suspected symptoms to seek care at TB-designated medical institutions, and enhancing public awareness of active case finding; in parallel, active TB screening should be expanded in priority populations and key settings, and a routine monitoring and early-warning system for high-risk groups should be established to reduce patient delay.

## Data Availability

All data generated or analysed during this study are included in this article.
Further enquiries can be directed to the corresponding author.

## Acknowledgements

We would like to acknowledge the hard and dedicated work of all the staff that implemented the intervention and evaluation components of the study

## Authors’ contributions

Conception and design of the research: He Fukui, Wang Xin.Acquisition of data: He Fukui, Wang Kaihao.

Analysis and interpretation of the data: He Fukui, Wang Xin, Wang Kaihao.Statistical analysis: He Fukui, Wang Xin. Writing of the manuscript: He Fukui.Critical revision of the manuscript for intellectual content: He Fukui.All authors read and approved the final draft.

## Availability of data and materials

All data generated or analysed during this study are included in this article. Further enquiries can be directed to the corresponding author.

## Declarations

### Ethics approval and consent to participate

I confirm that I have read the editorial policy page.This study was approved by the Ethics Committee of theTB-designated hospital in Huyanghe City.This study was conducted in accordance with the Declaration of Helsinki.All participants provided written informed consent.

### Consent for publication

Not applicable.

## References

[1] Organization World Health. Global tuberculosis report 2025 [M]. Geneva: World Health Organization.

[2] Hao Dongqing, Li Tao, Xu Caihong. Influencing factors associated with health-care seeking delay and diagnosis delay of pulmonary tuberculosis patients in western China, 2020[J]. Disease Surveillance, 2023, 38(11): 1294–1300.

[3] Technical Guidance Group of the Fifth National TB Epidemiological Survey;The Office of the Fifth National TB Epidemiological Survey. The fifth national tuberculosis epidemiological survey in 2010[J]. Chinese Journal of Antituberculosis, 2012, 34(8): 485–508.

[4] Getnet Fentabil, Demissie Meaza, Assefa Nega, et al. Delay in diagnosis of pulmonary tuberculosis in low-and middle-income settings: systematic review and meta-analysis [J]. BMC Pulmonary Medicine, 2017, 17(1): 202.

[5] Liu Kui, Ge Rui, Luo Dan, et al. Delay analysis of pulmonary tuberculosis in the eastern coastal county of China from 2010 to 2021: evidence from two surveillance systems [J]. Frontiers in Public Health, 2023, 11: 1233637.

[6] Ji Haoqiang, Xu Jia, Wu Ruiheng, et al. Cut-off points of treatment delay to predict poor outcomes among new pulmonary tuberculosis cases in Dalian, China: a cohort study [J]. Infection and Drug Resistance, 2021: 5521–5530.

[7] Subbaraman Ramnath, Nathavitharana Ruvandhi R, Mayer Kenneth H, et al. Constructing care cascades for active tuberculosis: a strategy for program monitoring and identifying gaps in quality of care [J]. PLoS medicine, 2019, 16(2): e1002754.

[8] Amare D., Alene K. A., Ambaw F. Effect of integrating traditional and modern healthcare systems on tuberculosis case detection in Ethiopia: a cluster randomized controlled study [J]. Infect Dis Poverty, 2025, 14(1): 16.

[9] Paziliya Aini, Yipaer Aiheiti, ZHAO Yuhua, Muhebuli Nuermaimaiti, WANG Rui, Gulijiamali Rexiding, LIU Nianqiang. Epidemiological characteristics of reported pulmonary tuberculosis in Xinjiang Uygur Autonomous Region, 2005-2023. China Tropical Medicine. 2025, 25(12): 1533–1539

[10] Chang Caiyi, Ma Xiaoling, Dong Gaimai, et al. Current status and influencing factors of diagnostic delay among elderly pulmonary tuberculosis patients in the Xinjiang Production and Construction Corps[J]. Bulletin of Disease Control and Prevention (China)**, 2025, 40(06): 23–27.

[11] Hu Yi, Xu Biao, Zhao Qi. Health-care seeking behavior and influencing factors of access to health care among outpatients with chronic cough in county hospitals in rural northern Jiangsu, China[J]. Chinese Journal of Epidemiology, 2004, 25(8): 650–654..

[12] Han Yang, Niu Xiaobin, Peng Ailing, et al. Health-care seeking delay among tuberculosis patients in Huaibei City, Anhui Province, 2017-2021 and its influencing factors[J]. Shanghai Journal of Preventive Medicine, 2023, 35(08): 758–763

[13] Cheng Shiming, Wang Ni, Zhou Lin, et al. Exploration of indicators related to different types of delay among newly diagnosed smear-positive pulmonary tuberculosis patients[J]. Chinese Journal of Antituberculosis, 2011, 33(10): 633–636.

[14] Li-Juan FU, Ye-Sheng Wang, Wen-Long Zhu, et al. Consultation delay and influencing factors among pulmonary tuberculosis patients in Huzhou City from 2008 to 2018 [J]. Chinese Journal of Disease Control & Prevention, 2021, 25(2): 235–239.

[15] Hui Chen, Yin-Yin Xia, Can-You Zhang, et al. Epidemic trends and characteristics of pulmonary tuberculosis in students in China from 2014 to 2018 [J]. Chinese Journal of Antituberculosis, 2019, 41(6): 662.

[16] Alene Muluneh, Assemie Moges Agazhe, Yismaw Leltework, et al. Patient delay in the diagnosis of tuberculosis in Ethiopia: a systematic review and meta-analysis [J]. BMC infectious diseases, 2020, 20(1): 797.

[17] Li Deyang, Su Deqi, Zhang Weisheng, et al. Influencing factors of health-care seeking delay, diagnosis delay, and detection delay among pulmonary tuberculosis patients in Urumqi City[J]. Preventive Medicine, 2020, 32(11): 1150–1154

[18] Liang Da. Analysis of influencing factors of health-care seeking delay among pulmonary tuberculosis patients in Qinghai Province and a Bayesian network modeling study[D]. Qinghai University, 2022.

[19] Li Wenhui. Spatiotemporal clustering patterns and influencing factors of pulmonary tuberculosis incidence in Dongguan City[D]. Guangdong Pharmaceutical University, 2021.

[20] Shu Qi, Shen Li, Yang Zhenyuan, et al. Health-care seeking delay among pulmonary tuberculosis patients in Jinshan District, Shanghai, 2014–2018 and its influencing factors[J]. Preventive Medicine Tribune, 2019, 25(07): 494-495, 498

[21] Li Xia, Zhang Jia, Ma Guorong, et al. Analysis of health-care seeking among tuberculosis patients in Yinchuan City, 2005–2017[J]. Preventive Medicine Tribune, 2021, 27(05): 321-324, 329

[22] Yu Nan, Wei Yunfang. Medical care seeking delay and related factors in pulmonary tuberculosis patients in Chaoyang district, Beijing, 2014–2020 [J]. Disease Surveillance, 2022, 37(1): 92–96.

[23] Wei Jianjun, Zeng Lingxia. Health-care seeking delay among pulmonary tuberculosis patients in Hanzhong City, Shaanxi Province, 2014–2017 and its influencing factors[J]. Journal of Public Health and Preventive Medicine, 2018, 29(5): 55–58

[24] Sun Minglei, Wu Qunhong, Guan Li, et al. Health-care seeking delay among tuberculosis patients in Heilongjiang Province in 2004 and its influencing factors[J]. Journal of Tuberculosis and Lung Disease, 2020, 1(4): 270–275

[25] Batbayar Batmunkh, Kariya Tetsuyoshi, Boldoo Tsolmon, et al. Patient delay and health system delay of patients with newly diagnosed pulmonary tuberculosis in Mongolia, 2016–2017 [J]. Nagoya journal of medical science, 2022, 84(2): 339.

[26] Guo Jing, Feng Yin-Ping, Liu Zhong-Da, et al. Analysis of factors influencing patient delay by patients with pulmonary tuberculosis in Lishui City, Zhejiang Province [J]. BMC Pulmonary Medicine, 2023, 23(1): 264.

[27] Bojovic Olivera, Medenica Milic, Zivkovic Danko, et al. Factors associated with patient and health system delays in diagnosis and treatment of tuberculosis in Montenegro, 2015–2016 [J]. PLoS One, 2018, 13(3): e0193997.

[28] Auer Christian, Kiefer Sabine, Zuske Meike, et al. Health-seeking behaviour and treatment delay in patients with pulmonary tuberculosis in Switzerland: some slip through the net [J]. Swiss medical weekly, 2018, 148.

[29] Cox Helen S, Mbhele Slindile, Mohess Neisha, et al. Impact of Xpert MTB/RIF for TB diagnosis in a primary care clinic with high TB and HIV prevalence in South Africa: a pragmatic randomised trial [J]. PLoS medicine, 2014, 11(11): e1001760.

[30] Telisinghe L, Ruperez M, Amofa-Sekyi M, et al. Does tuberculosis screening improve individual outcomes? A systematic review [J]. EClinical Medicine, 2021, 40.

[31] Wu Z, Chen J, Xia Z, et al. Impact of the COVID-19 pandemic on the detection of TB in Shanghai, China [J]. Int J Tuberc Lung Dis, 2020, 24(10): 1122–1124.

[32] Liu Qiao, Lu Peng, Shen Ye, et al. Collateral impact of the coronavirus disease 2019 (COVID-19) pandemic on tuberculosis control in Jiangsu Province, China [J]. Clinical infectious diseases, 2021, 73(3): 542–544.

[33] Xia Yinyin, Huang Fei, Chen Hui, et al. The impact of COVID-19 on tuberculosis patients’ behavior of seeking medical care—China, 2020 [J]. China CDC Weekly, 2021, 3(26): 553.

[34] Chen H., Zhang K. Insight into the impact of the COVID-19 epidemic on tuberculosis burden in China [J]. Eur Respir J, 2020, 56(3).

[35] Fei Huang, Yinyin Xia, Hui Chen, et al. The impact of the COVID-19 epidemic on tuberculosis control in China [J]. The Lancet Regional Health–Western Pacific, 2020, 3.

[36] Demissie Meaza, Lindtjorn Bernt, Berhane Yemane. Patient and health service delay in the diagnosis of pulmonary tuberculosis in Ethiopia [J]. BMC Public Health, 2002, 2(1): 23.

[37] Enkhbat Shagdarsurengiin, Toyota Makoto, Yasuda Nobufumi, et al. Differing influence on delays in the case-finding process for tuberculosis between general physicians and specialists in Mongolia [J]. Journal of epidemiology, 1997, 7(2): 93–98.

[38] Xu J, Luo P, He XX. Analysis of characteristics and diagnosis delay of 629 elderly tuberculosis patients [J]. J Tuberc Lung Dis, 2021, 2(3): 216–222.

[39] Guan Fy Chen ZH, Li WH, et al. Characteristics analysis of diagnosis delay among elderly tuberculosis patients in Dongguan City from 2009 to 201?8 [J]. J Tuberc Lung Dis, 2021, 2(3): 243–250.

[40] Xu Jia. Analysis of health-care seeking delay and diagnostic delay and their influencing factors among migrant pulmonary tuberculosis patients[D]. Dalian Medical University, 2022

[41] Storla D. G., Yimer S., Bjune G. A. A systematic review of delay in the diagnosis and treatment of tuberculosis [J]. BMC Public Health, 2008, 8: 15.

[42] Yang W. T., Gounder C. R., Akande T., et al. Barriers and delays in tuberculosis diagnosis and treatment services: does gender matter? [J]. Tuberc Res Treat, 2014, 2014: 461935.

[43] Krishnan L., Akande T., Shankar A. V., et al. Gender-related barriers and delays in accessing tuberculosis diagnostic and treatment services: a systematic review of qualitative studies [J]. Tuberc Res Treat, 2014, 2014: 215059.

[44] Bonadonna L. V., Saunders M. J., Zegarra R., et al. Why wait? The social determinants underlying tuberculosis diagnostic delay [J]. PLoS One, 2017, 12(9): e0185018.

[45] Feng J. Y., Huang S. F., Ting W. Y., et al. Gender differences in treatment outcomes of tuberculosis patients in Taiwan: a prospective observational study [J]. Clin Microbiol Infect, 2012, 18(9): E331–337.

[46] Fu Lijuan, Wang Yesheng, Zhu Wenlong, et al. Health-care seeking delay among pulmonary tuberculosis patients in Huzhou City, 2008–2018 and its influencing factors[J]. Chinese Journal of Disease Control & Prevention, 2021, 25(2): 235–239

[47] Chen HG, Liu M, Jiang SW, et al. Impact of diabetes on diagnostic delay for pulmonary tuberculosis in Beijing [J]. The International journal of tuberculosis and lung disease, 2014, 18(3): 267–271.

[48] Jeon Christie Y, Murray Megan B. Diabetes mellitus increases the risk of active tuberculosis: a systematic review of 13 observational studies [J]. PLoS medicine, 2008, 5(7): e152.

[49] Fetensa Getahun, Wirtu Desalegn, Etana Belachew, et al. Magnitude and determinants of delay in diagnosis of tuberculosis patients in Ethiopia: a systematic review and meta-analysis: 2020 [J]. Archives of Public Health, 2022, 80(1): 78.

[50] Storla Dag Gundersen, Yimer Solomon, Bjune Gunnar Aksel. A systematic review of delay in the diagnosis and treatment of tuberculosis [J]. BMC Public Health, 2008, 8(1): 15.

